# Functional connectivity of reward and cognitive control networks predicts body mass index but not binge eating disorder in women

**DOI:** 10.64898/2026.11.10.26367718

**Authors:** Antonia S. Schlaich, María Berjano Torrado, Anne Kühnel, Jennifer Svaldi, Nils B. Kroemer

## Abstract

**Background:** Binge eating disorder (BED) is associated with obesity as well as behavioral and neurobiological changes in the reward and cognitive control systems of the brain. To better dissociate neural mechanisms of pathological eating behavior, functional connectivity (FC) analyses have shown promise. However, there is currently no replicable signature of BED derived from fMRI.

**Methods:** We investigated FC in 61 women with varying symptoms of binge eating. The three groups included 21 women with BED, 20 women with subsyndromal BED (subBED), and 20 controls without binge eating (noBE) matched in body mass index (BMI) and age. To identify neural signatures of BED and BMI, we used seed-based FC “fingerprints” (nucleus accumbens, NAcc, for reward and dorsolateral prefrontal cortex, dlPFC, for cognitive control) to predict group or BMI based on resting state and task-based (food bidding) FC. Furthermore, we explored spatial patterns of FC to quantify associations with BED and BMI.

**Results:** Both seeds of the reward and cognitive control networks robustly predicted BMI (*r*s = .33 – .46). The dlPFC seed predicted BMI across conditions (resting state: *p=*.002; task: *p=*.012) while the NAcc seed only predicted BMI based on task-based FC (*p=*.007). In contrast, FC failed to classify binge eating groups better than chance.

**Conclusions:** FC within reward and cognitive control networks primarily reflect obesity-related changes. Despite pronounced differences in eating behavior, BED could not be reliably dissociated from subBED or noBE using FC fingerprints. These results highlight challenges in identifying BED-specific functional markers.

**Graphical Abstract:** 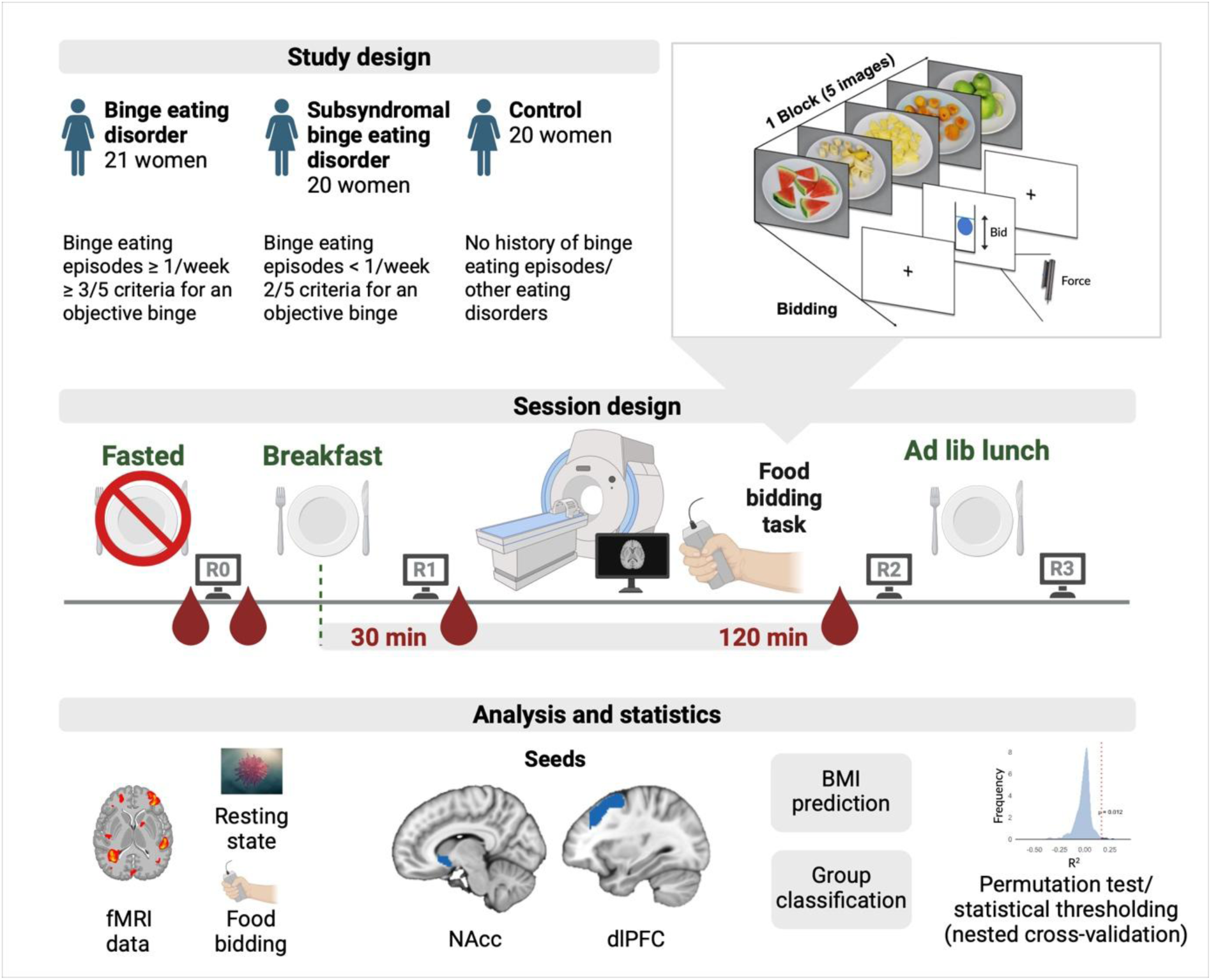

Created with BioRender.com.

## Introduction

Binge eating disorder (BED) is the most prevalent eating disorder (1) with a lifetime prevalence of 0.6–1.8% in women and 0.3–0.7% in men (2). BED is characterized by a lack of control over eating, leading to the consumption of unusually large amounts of food in a short time (3). Since the definition of BED excludes compensation of binge eating episodes, it is typically associated with overweight or obesity (4). Therefore, patients with BED also show an increased risk for physical comorbidities associated with obesity including diabetes, metabolic syndrome, and cardiovascular diseases (5). Moreover, BED is associated with an increased incidence of mental disorders such as depression, anxiety (4), attention deficit hyperactivity disorder (6), or addiction (7, 8) that are characterized by higher levels of impulsivity (9, 10). Despite the known comorbidity of overweight/obesity and BED(4), discerning changes in brain function remains challenging. Elucidating differences in brain function that relate specifically to BED may facilitate improved treatment options for BED beyond psychotherapy (11).

On a neurobiological level, BED has been associated with altered function in the reward and cognitive control systems. Individuals with BED show increased activation of reward-related regions during the presentation of food stimuli in passive food-viewing tasks (12) and during food consumption in active food-tasting tasks (13) compared to matched controls. However, other studies found decreased neural activity in response to high-energy foods that may be consistent with those consumed during a binge eating episode (14). This pattern is consistent with the notion that greater severity is associated with heightened anticipatory reward processing (“wanting”), alongside relatively diminished consummatory reward processing (“liking”), specifically in response to food rewards (15).

On a behavioral level, rash-spontaneous behavior is more frequent (16) and cognitive control is decreased in patients with BED (17). Reduced cognitive control in patients with BED is reflected in lower functional connectivity (FC), lower fiber tract integrity, and higher structural connectivity between the frontal cortex and the striatum (18). Accordingly, resting state FC between the middle frontal gyrus (midFG) and the insula is lower in participants with BED (19). Similarly, FC between cognitive control (dorsolateral prefrontal cortex, dlPFC) and reward/salience networks is reduced in pre-adolescent children (amygdala and orbitofrontal cortex, (20)) and adults(striatum, (21)) with BED. Taken together, BED is characterized by altered connectivity between cognitive control and reward networks, which may contribute to the loss of control over eating (21). Despite the growing number of studies examining FC in BED, it is not known whether presenting disorder-related stimuli (e.g., food) in a motivational context, such as bidding, would improve the accuracy of prediction and classification.

In contrast to BED, FC alterations in overweight and obesity have been more extensively investigated (22). Obesity has been associated with disrupted FC within large-scale brain networks, including the salience network, the default mode network (DMN), and the frontoparietal network (23). These alterations have been linked to heightened incentive salience of food cues, impaired cognitive control over eating, and increased interoceptive awareness of hunger signals (23, 24). Specifically, resting state FC of the anterior cingulate cortex (ACC) is reduced in obesity, whereas increased connectivity of the posterior cingulate cortex (PCC) and precuneus has been observed, indicating altered DMN organization (25). Additionally, altered resting state FC of the NAcc has been reported in individuals with obesity (26) and differences in FC with the primary ingestive cortex and the ventromedial prefrontal cortex predict changes in eating behavior in patients with depression (27), highlighting the central role of reward-related circuits. Although similar FC alterations have been observed during food-related tasks, with reduced DMN FC during food picture viewing (28) and increased FC within the salience network during food-related working memory tasks (29), many studies have not directly compared resting and tasks states.

Often, neuroimaging studies investigating obesity do not control for binge eating symptoms, potentially conflating associations of body weight with pathological eating (30, 31). Since BED is strongly associated with obesity, it is still unclear whether differences in brain function are largely driven by obesity or if specific signatures indicative of pathological eating can be isolated. To address this gap and disentangle shared and distinct FC alterations between BED and obesity, we examined FC in a sample of 61 overweight or obese women, including 21 individuals diagnosed with BED, 20 individuals exhibiting current binge eating behavior without meeting the diagnosis criteria for BED (i.e., subsyndromal BED, subBED), and 20 weight-matched controls without binge eating (noBE). Crucially, we investigated both resting state and task-based FC during a food bidding task, capitalizing on FC in response to relevant stimuli, which has been suggested to improve brain-behavior associations (32). To capture the functional relevance of reward and cognitive control processes, we used seed-to-ROI (whole brain) FC profiles from the NAcc and dlPFC to predict BED and BMI.

## Methods

### Participants

In total, 61 females divided into three groups participated in the study (*M*_age_ = 39.7 years ± 13.2, range 21 and 64 years; *M*_BMI_ = 31.5 kg/m^2^ ± 7.1). The first group included individuals diagnosed with BED. The second group were classified as subBED because they reported objective binge-eating episodes during the preceding 12 months but failed to meet the frequency criterion for a diagnosis of BED, met only two instead of three out of five existing criteria for an objective binge, or lacked marked distress regarding binge eating. The third group of participants reported having no lifetime history of subjective and objective binge eating or other eating disorders (noBE). The noBE group was matched to participants with BED according to age and BMI.

Before the first session, all participants underwent standardized phone screenings to verify eligibility and assign everyone to one of the three study groups. To assess BED symptoms, we used the German version of the Eating Disorder Examination (EDE) (33), while the Structured Clinical Interview for DSM IV (SCID) (34) was used to screen for other mental disorders. Since BED is often associated with symptoms of depression or other mental disorders (1, 35), participants with depression were also included if their antidepressant medication was stable and they did not report acute suicidal tendencies. We excluded individuals who reported other eating disorders, co-occurring psychotic or bipolar disorder, severe alcohol/substance dependence within the past six months, medical disorders that would affect weight or the ability to participate, or insufficient German language skills. Additionally, we screened for exclusion criteria for MRI.

The study was approved by the ethics committee (i.e., the institutional review board) of the University of Tübingen (393/2017BO2) and conducted in accordance with the Declaration of Helsinki, as revised in 2008. Informed consent was obtained from all participants prior to taking part in the experiment. Participants received either monetary compensation (∼90€) or partial course credit for their participation. The exact amount varied based on their performance during the tasks.

### Experimental procedure

The study comprised an online assessment (36, 37) and two sessions of laboratory assessments: a behavioral session and an fMRI session. For the fMRI session, participants were asked to fast overnight (i.e., >8 h before the session) and we analyzed fasting levels of glucose, insulin, triglycerides, and ghrelin. We also collected repeated ratings of mood and metabolic state (38, 39). Thereafter, participants received a standardized breakfast, which ensured that they entered the MRI in a non-fasting state to increase inter-individual differences in brain responses to food. The fMRI measurement started with an anatomical scan, followed by resting state using the Inscapes movie (40), an effort allocation task (not analyzed here; (36, 41)) and the food bidding task. The fMRI session lasted about 100 min. By the end of the session, participants received the previously earned snacks and money.

### Food bidding task

In the food bidding task, participants were presented with standardized pictures of food and control objects (e.g., stationery;(42)) and asked to bid as much as they wanted to receive the presented objects by exerting physical effort. Food items were presented in 12 blocks (5 images per block) of four categories (high-calorie savory, high-calorie sweet, low-calorie savory, low-calorie sweet), and images were shown twice. Prior to each block, a fixation cross was displayed. At the end of the cue reactivity phase, participants were asked to indicate how much physical effort they were willing to exert to gain access to a buffet including the items shown previously. By pressing the grip force device, a blue ball shown in a vertical tube on the screen could be lifted, with the height of the ball being proportional to the force participants applied. The force was determined relative to each participant’s maximum force that had previously been recorded in a training phase of an effort allocation task (41, 43, 44). Participants were asked to keep the ball at the height corresponding to the effort they were willing to exert for about five seconds during the bidding phase. The task took 14 min.

### fMRI data acquisition

Structural and functional MRI data were collected using a Siemens 3 T PRISMA whole-body MR scanner (Siemens Medical Solutions, Erlangen, Germany) with a 64-channel RF head receiver coil. Details of the imaging sequences were collected following the COBIDAS consortium guidelines (45). T1 weighted structural images were measured using an MPRAGE sequence with 176 sagittal slices covering the whole brain, flip angle = 9°, matrix size = 256 × 256 and voxel size = 1 × 1 × 1 mm³. Field maps were acquired by a Siemens gradient echo field map sequence with a short (5.19 ms) and a long (7.65 ms) echo time (TE), which resulted in a TE difference of 2.46 ms. fMRI data consisting of 10 minutes resting state (430 volumes) and the food bidding task (594 volumes) was acquired as T2*-weighted gradient echo-planar images (EPIs) using a multiband factor of 4, 68 axial slices with an interleaved slice order covering the whole brain including brainstem, repetition time (TR) = 1.4 s, TE = 30 ms, flip angle = 65°, 110 × 110 matrix, field of view = 220 × 220 mm² and voxel size = 2 × 2 × 2 mm³. During resting state fMRI, we showed the Inscapes video without audio to improve compliance while minimizing cognitive load and motion (40).

### fMRI data preprocessing

Preprocessing of resting state fMRI data was performed using fMRIPrep 20.1.1 (46), based on Nipype 1.4.1 (47, 48). This included intensity non-uniformity correction, skull stripping, brain tissue segmentation, brain surface reconstruction, and volume-based spatial normalization. Distortion correction and co-registration to T1-weighted images were also applied. Head-motion parameters were estimated, and BOLD runs were slice-time corrected. Further preprocessing, including smoothing (Gaussian kernel of 6 mm full-width half maximum), bandpass filtering (0.01 Hz and 0.1 Hz), and denoising (six movement parameters and their derivative, 5 components of white matter and cerebrospinal fluid signal each, and outlier scans, see SI), was performed using CONN (49) with default settings and SPM 12 (50).

### Data analysis

#### Seed-based functional connectivity

To compute seed-based FC, we estimated seed-to-ROI connectivity vectors for each participant for both resting and task states using CONN (49). Fisher-*z* transformed correlation coefficients between each seed region and all other ROIs represented FC strength. To capture FC related to cognitive control and reward processing, we selected two seed regions as defined by the Harvard-Oxford brain atlas, namely the NAcc (left and right) and the midFG (left and right), corresponding to the dlPFC (Fig. 1c). To compare our results with previous work, we also included the ACC as a seed, which has shown distinct FC profiles in obesity (25). To reduce the number of features (due to the moderate sample size), we averaged FC values for regions for both hemispheres. Hence, we used seed-to-ROI FC vectors (fingerprints) with 81 connections for each seed for prediction and classification.

**Figure 1:**
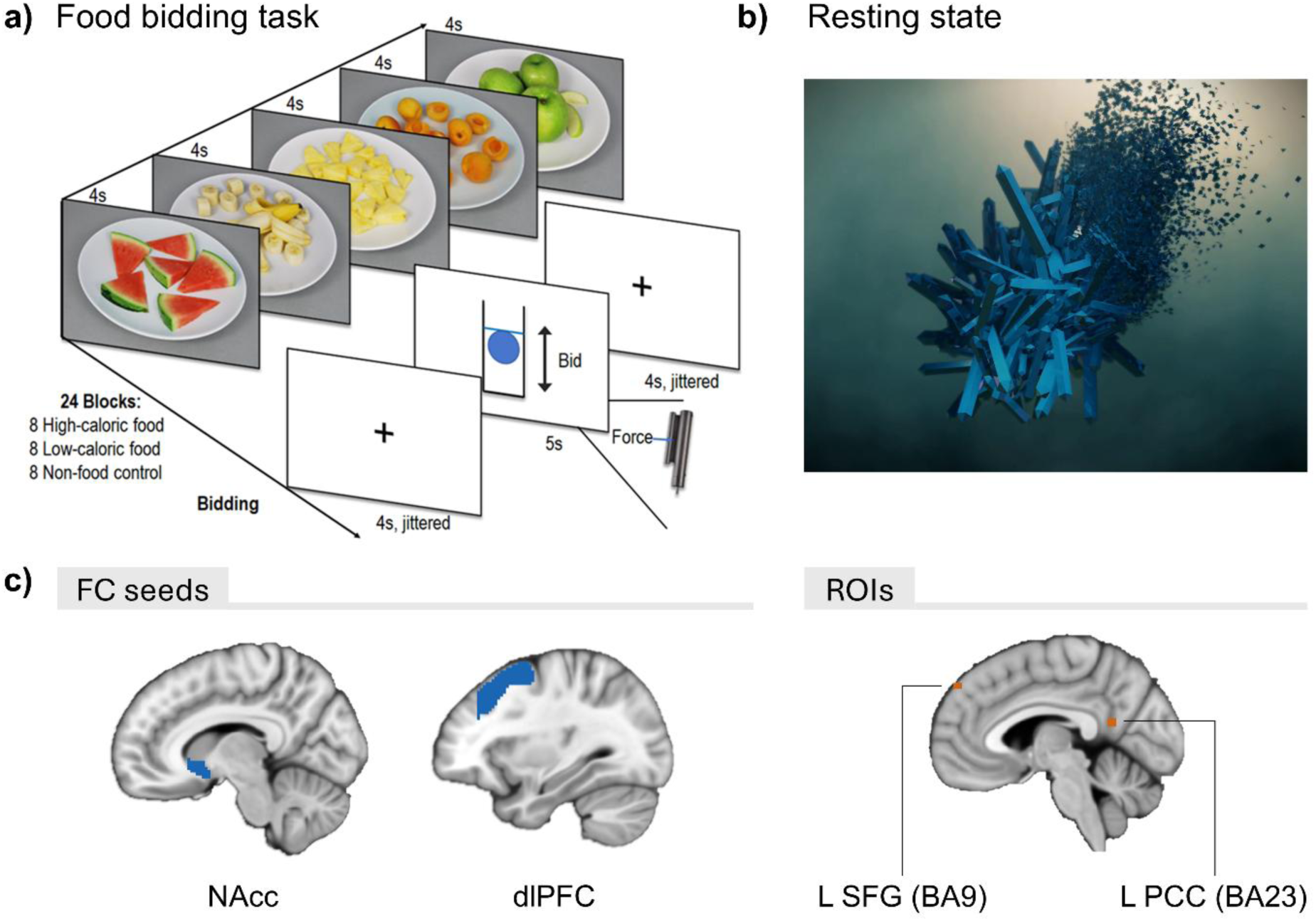
Overview of experimental tasks and connectivity regions of interest. a) Illustration of the food bidding task performed in the scanner. Each block starts with 4 s presentation of each picture for that block. Then, after a fixation cross is shown for 4 s (jittered), participants indicate their willingness to work for the presented food by pressing a grip force device during 5 s. The height of a blue ball inside a vertical tube depicted on the screen represents the force applied by the participants relative to their maximum force. b) Snapshot from *Inscapes*. *Inscapes* is a computer-generated animation with a fixed camera angle and no scene cuts that was shown during acquisition of resting state data (40). c) FC seeds and regions of interest (ROIs). The NAcc seed was defined using the Harvard-Oxford atlas. As there are no specific dlPFC parcellations in said atlas, we used the regions corresponding to the middle frontal gyrus (midFG). The lower right panel shows the ROIs reported by Haynos et al. (2021) that showed lower NAcc connectivity in BED vs. control groups. These ROIs were generated to replicate seed-based NAcc differences.

#### Connectome-based prediction of BED groups and BMI

To classify group differences and predict BMI based on FC, we used seed-to-ROI FC vectors from resting and task states.

##### BED group classification

To evaluate the discriminative capacity of FCs among distinct BED groups, we used a multiclass classification approach. The classification model consisted of a one-versus-all 3-class error-correcting output codes (ECOC) model with ensembles of decision trees as binary learners (100 ensemble learning cycles). This method involves training a separate binary classifier for each subset. The decision boundaries were defined by the ensemble learning method GentleBoost, a more stable and robust version of AdaBoost (51), which combines the predictions of multiple weak learners (decision trees). To guard against overfitting, we evaluated the model fit using 10-fold cross-validation (52). To assess the statistical significance of the predictive model accuracy, we performed permutation tests using a baseline model that included only age and BMI as confounders (1000 iterations; outcome was shuffled with confounders to keep their correlation structure; (53)). We fitted the model separately to resting and task state data for each seed region (NAcc, dlPFC, and ACC).

##### BMI prediction model

To predict BMI, we used seed-to-ROI FCs fingerprints and elastic net regression (lasso, preset alpha = 0.5) with nested 10-fold cross-validation (inner loop folds = 10; outer loop folds = 10) (see 53, 54). We iterated this process 20 times to select a typical model (i.e., *R*² closest to the median). Statistical significance was determined by using permutation tests (1,000 iterations), including age and group (BED, subBED, or noBE) as control variables. We performed two BMI predictions for each seed region using the seed-to-ROI resting state FC and task FC data. To further characterize the associations of FC with BMI, we explored the relationship between BMI and FC patterns in seeds that provided significant BMI predictions. We extracted the weights of the regions that contributed to the BMI prediction to create β-weight maps for all seed regions and resting and task states.

#### Seed-based FC maps and group analyses

To characterize the spatial patterns of FC related to the prediction of BED or BMI, we used exploratory whole-brain seed-based FC maps from the NAcc, dlPFC, and ACC seeds. To test for group differences, we used the individual seed-based maps in SPM12 (50). We estimated factorial designs for each of the three seeds and included the three groups, BMI, and age as predictors. Contrasts of interest included positive and negative correlations with BMI, as well as comparisons between the BED and noBE groups, indicative of a linear increase from noBE to subBED and BED.

To replicate the findings of Haynos et al. (21), we tested regions that showed significant resting state FC differences between individuals with BED and noBE with NAcc as the seed. We generated the ROIs as spheres with 2 mm radius using MarsBaR within SPM (55) at the reported significant peaks with the following MNI coordinates: x = −6, y = 52, z = 43 (left superior frontal gyrus; L SFG, BA 9); and x = - 2, y = −54, z = 18 (left PCC, BA 23) and combined them into a single mask for small volume correction (SVC; Fig. 1c).

#### Statistical analyses and software

We preprocessed fMRI data using fMRIPrep 20.1.1 (46, 56), based on Nipype 1.4.1, CONN release 22.a (49), and SPM12 release 12.7771 (50). ROIs were created using MarsBaR. All statistical analyses were conducted with MATLAB R2022b or R 4.2.2 (57). We considered α ≤ .05 as significant. Data were visualized using Mango 4.1, ggplot2 (58), and cvms (https://github.com/ludvigolsen/cvms).

## Results

To investigate differences in FC patterns in BED and obesity, we performed a comprehensive analysis involving the prediction of BMI and BED status, as well as the characterization of the spatial FC patterns using a seed-to-ROI approach.

### Seed-to-ROI FC patterns do not differentiate BED

To assess whether FC patterns are indicative of the participants’ binge eating behavior (BED status), we applied a supervised classification model on seed-to-ROI FC data. Seeds were selected within the reward (NAcc) and cognitive control (dlPFC) networks, and both resting state and task-based FC data were evaluated (for the ACC, see SI). Across all seeds and modalities, the models did not predict participants’ BED status above chance (all *p*s > .106, Fig. 2; See Fig. S2 for ACC results).

**Figure 2.**
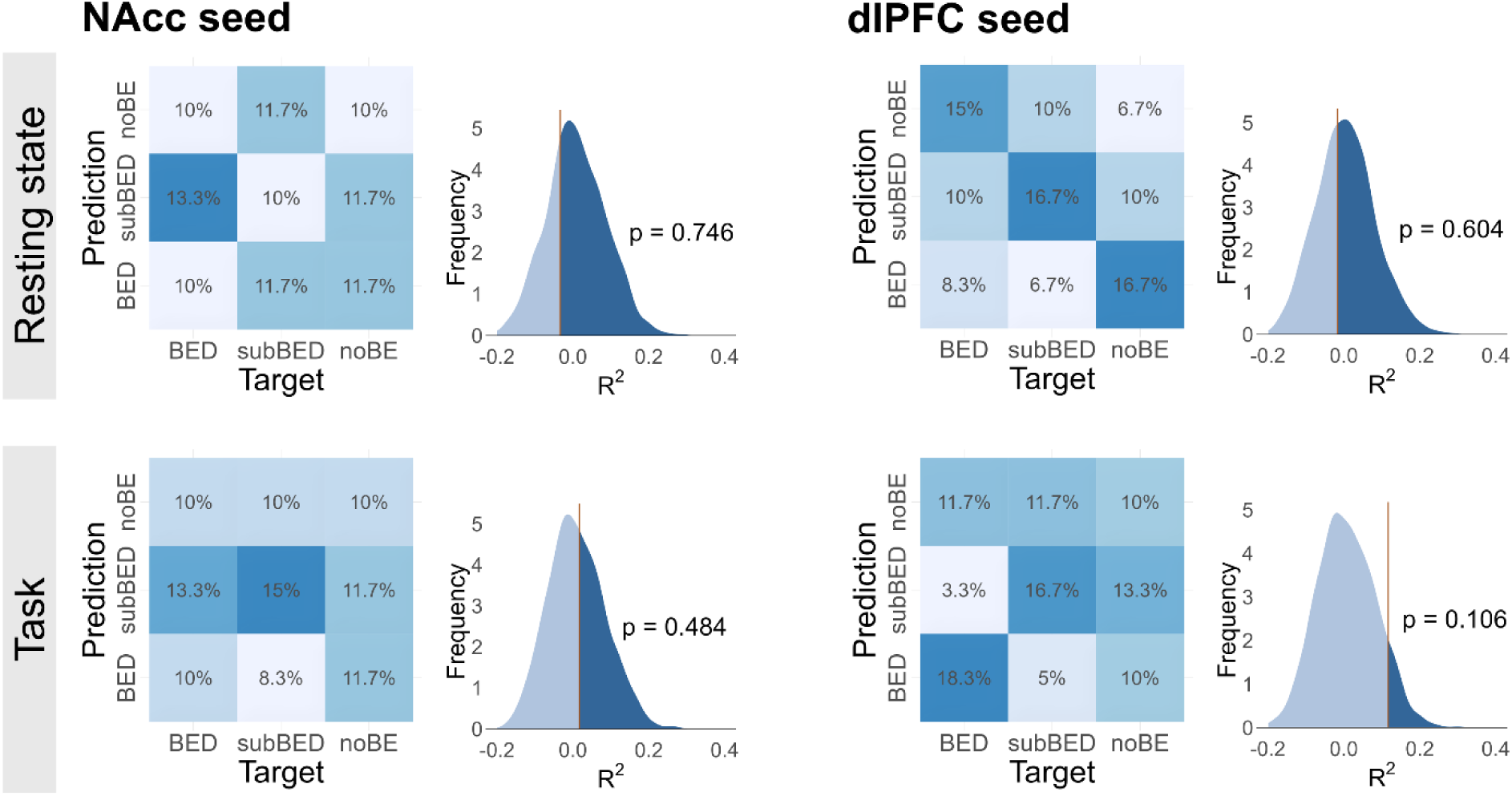
BED status cannot be classified better than chance based on seed-to-ROI FC for resting state and task fMRI. NAcc-to-ROI and dlPFC-to-ROI classification accuracy are shown for resting state and task conditions. For each seed and condition, a multiclass confusion matrix shows the normalized proportion of participants in each observed group (columns) classified into each predicted group (rows). Additionally, the distribution of the classification accuracy generated by permutation testing is shown, with a red vertical line representing the observed data test statistic. The median accuracy of the permutation test was used as a comparison baseline (1,000 iterations).

### FC differences between BED and control groups

Since the prediction of BED by multivariate FC fingerprints was unsuccessful across all selected seeds and conditions, we explored whether this approach missed specific changes in BED. First, we attempted to replicate the results reported by Haynos et al. (21), in which the NAcc showed lower resting FC with the left SFG and left PCC in BED v. control. We compared the NAcc-to-whole-brain maps of resting state FC between noBE vs. BED using small-volume correction for the reported ROIs. Contrary to our expectations, no significant group differences were observed in NAcc-SFG or NAcc-PCC connectivity (*t* = 0.00, *p* = 0.83, and *t* = −0.64, *p* = 0.88, respectively; Table S1). Moreover, while the participants belonging to the BED group in Haynos et al. (21) showed reduced NAcc-to-ROI FC compared to the control group, our results indicate that the BED group FC was slightly elevated relative to the noBE group when comparing the participant’s mean FC for each ROI (NAcc-to-SFG: *d* = −0.20; NAcc-to-PCC: *d* = −0.42, where the negative sign indicates that the noBE group had a lower mean FC than the BED group; Fig. 3). Additionally, we examined group differences in seed-to-voxel maps for both task and rest conditions for the NAcc, dlPFC, and ACC seed regions. No clusters survived whole-brain correction for multiple comparisons (see unthresholded maps in Figure S1 and neuroVault for use in future studies; https://identifiers.org/neurovault.collection:17852).

**Figure 3.**
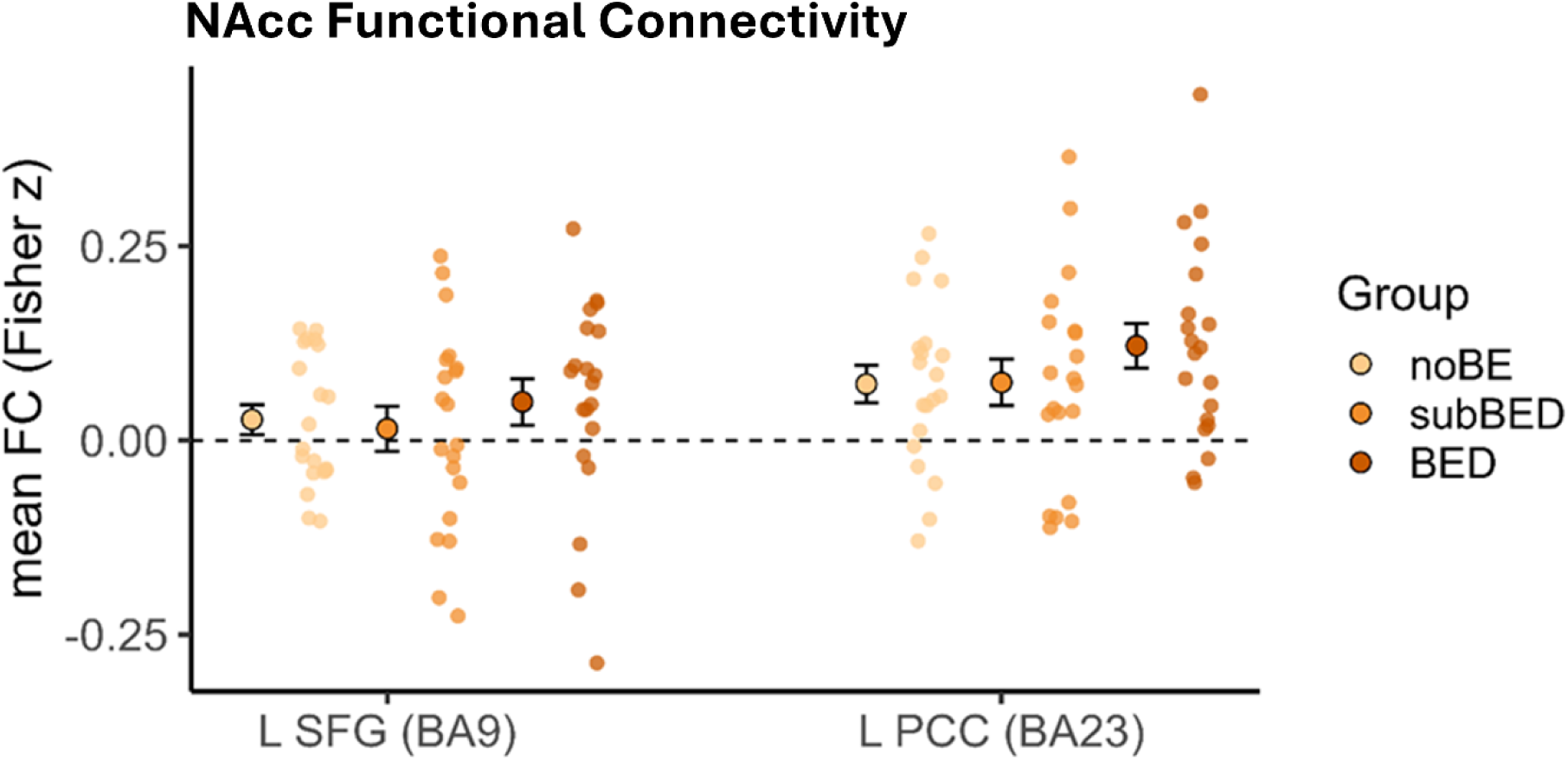
No significant differences in functional connectivity (FC) between NAcc and previously reported regions differenciating BED and control groups by Haynos et al. (2021). The NAcc FC is depicted for each ROI and BED status group. Small data points represent the mean FC per ROI of individual participants. Large points represent the mean FC for each group and the error bars represent the standard error of the mean (SEM).

### Seed-to-ROI FC patterns predict BMI

We next examined whether FC patterns were associated with participants’ BMI. Using seed-to-ROI FC from the resting state and task conditions, elastic net regression models significantly predicted BMI for multiple seeds. In the task condition, FC patterns of all ROIs predicted BMI (NAcc: *p* = 0.007, R^2^ = 0.199; dlPFC: *p =* 0.012, R^2^ = 0.166; ACC: *p* = 0.001, R^2^ = 0.258). Statistical significance was determined using permutation tests (1,000 iterations) with a baseline model that included age and BED groups as confounders. In contrast, resting state FC was only predictive of BMI from the dlPFC seed (*p* = 0.002, R^2^ = 0.243), while NAcc and ACC FC did not predict BMI (*p* = 0.69, R^2^ = −0.029; and *p* = .67, R^2^ = -.022, respectively; Fig. 4 and SI).

**Figure 4.**
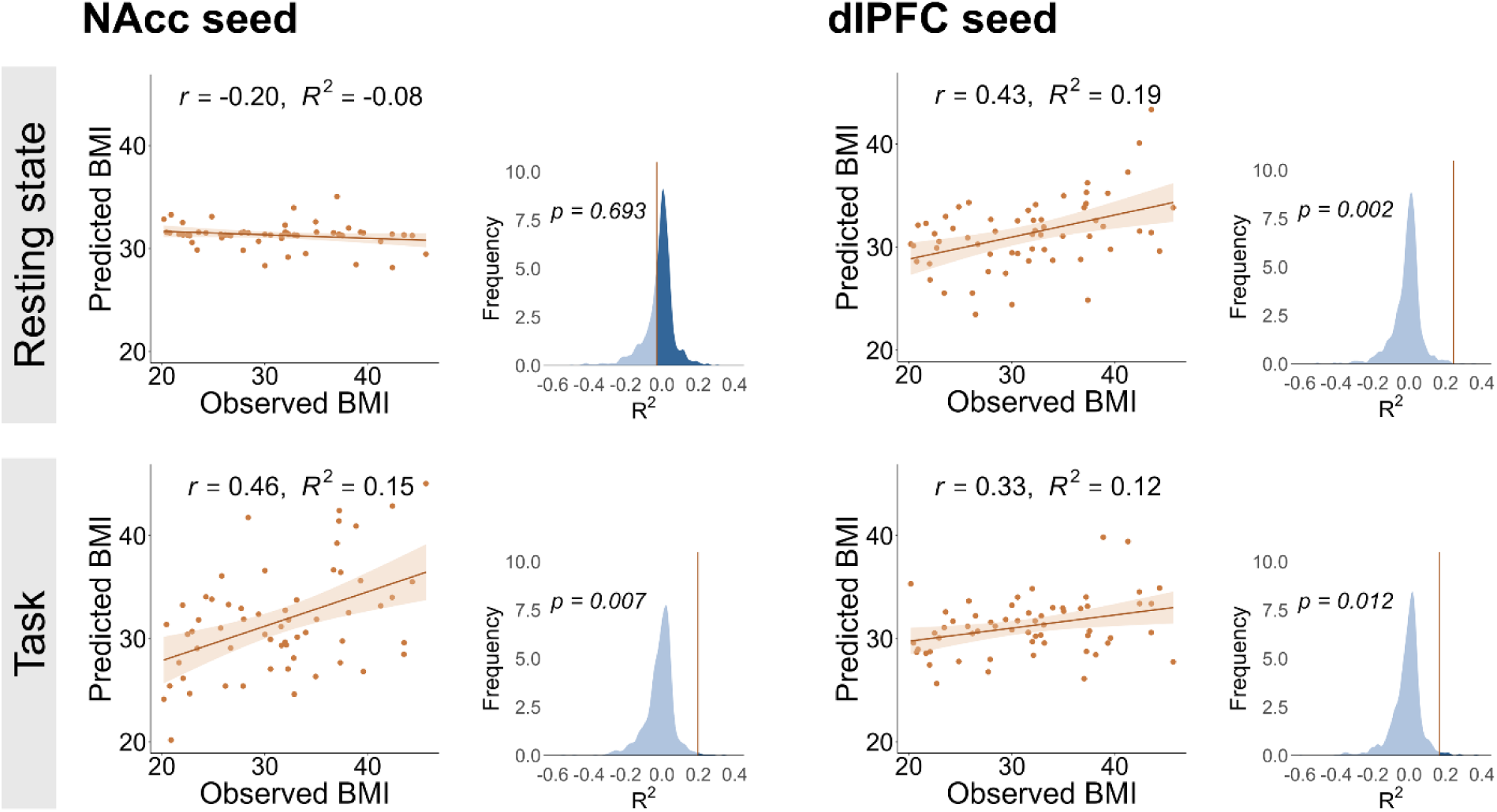
BMI can be successfully predicted based on seed-to-ROI FC for resting state and task fMRI. NAcc-to-ROI and dlPFC-to-ROI prediction accuracy are shown for resting state and task conditions. For each seed and condition, a scatter plot displays the relationship between observed BMI and predicted BMI, with fitted regression lines and shaded 95% confidence intervals. Additionally, a distribution of the coefficient of determination (*R*^2^ measure of goodness of fit) generated by permutation testing is shown, with the vertical red line representing the observed test statistic (1,000 iterations). For visualization, the permutation-derived R² values were centered by subtracting the median of the null distribution, such that zero corresponds to the median permutation performance. Negative values indicate prediction performance below the median of the null distribution, and positive values indicate prediction performance above the median of the null distribution.

To determine which connectivity patterns contributed to the significant BMI prediction, we examined the standardized elastic net regression coefficients, or β-weights, from the seed-to-ROI models that predicted BMI: task-based NAcc-to-ROI, resting-state dlPFC-to-ROI, and task-based dlPFC-to-ROI connectivity. Regarding the NAcc seed, frontal and dorsal striatum regions were among the strongest contributors to the BMI prediction, although with mixed signs (higher connectivity with the putamen and lower connectivity with the caudate). Other contributions were observed for subcortical regions implicated in homeostatic and autonomic regulation, including higher hypothalamic connectivity and decreased connectivity with the nucleus tractus solitarius (NTS) and brainstem. For the dlPFC models, connectivity with the caudate contributed positively to BMI prediction in both resting-state and task conditions, together with frontal cortical regions, including the inferior frontal gyrus (Figure 5).

**Figure 5.**
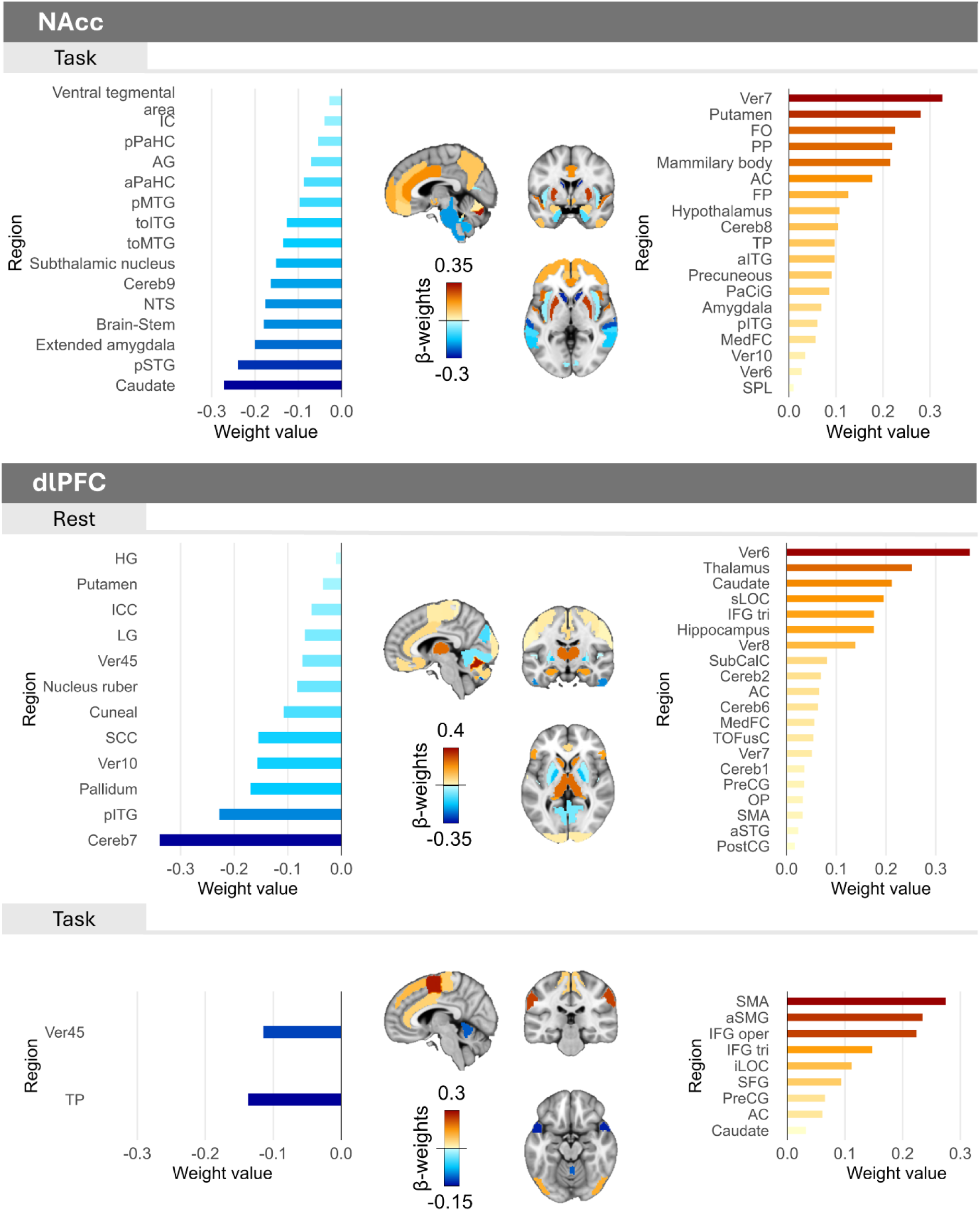
BMI prediction is supported by increased frontal, striatal, and somatomotor connectivity. Standardized β-weights for FC features retained in at least 80% of cross-validation iterations are shown for each seed region and condition. Positive weight values (warm colors) indicate features contributing positively to BMI prediction, whereas negative weight values (cold colors) indicate features contributing negatively. Across models, the strongest positive weights were predominantly observed within frontal and dorsal striatal, and somatomotor regions, with additional cerebellar, and subcortical structures. Seed regions were excluded from their respective predictor sets. *AC = Anterior cingulate cortex; AG = Angular gyrus; aITG = Anterior inferior temporal gyrus; aPaHC = Anterior parahippocampal gyrus; aSMG = Anterior supramarginal gyrus; aSTG = Anterior superior temporal gyrus; Cereb1 = Lobule I of the cerebellum; Cereb2 = Lobule II of the cerebellum; Cereb6 = Lobule VI of the cerebellum; Cereb7 = Lobule VII of the cerebellum; Cereb8 = Lobule VIII of the cerebellum; Cereb9 = Lobule IX of the cerebellum; dlPFC = Dorsolateral prefrontal cortex; FO = Frontal operculum cortex; FP = Frontal pole; HG = Heschl’s gyrus; IC = Insular cortex; ICC = Intracalcarine cortex; IFG oper = Inferior frontal gyrus, pars opercularis; IFG tri = Inferior frontal gyrus, pars triangularis; iLOC = Inferior lateral occipital cortex; LG = Lingual gyrus; MedFC = Medial frontal cortex; NAcc = Nucleus accumbens; NTS = Nucleus of the solitary tract; OP = Occipital pole; PaCiG = Paracingulate gyrus; pITG = Posterior inferior temporal gyrus; pMTG = Posterior middle temporal gyrus; pPaHC = Posterior parahippocampal gyrus; pSTG = Posterior superior temporal gyrus; PP = Planum polare; PreCG = Precentral gyrus; PostCG = Postcentral gyrus; sLOC = Superior lateral occipital cortex; SCC = Supracalcarine cortex; SFG = Superior frontal gyrus; SMA = Supplementary motor area; SPL = Superior parietal lobule; SubCalC = Subcallosal cortex; toITG = Temporo-occipital inferior temporal gyrus; toMTG = Temporo-occipital middle temporal gyrus; TOFusC = Temporo-occipital fusiform cortex; TP = Temporal pole; Ver6 = Vermis 6; Ver7 = Vermis 7; Ver8 = Vermis 8; Ver10 = Vermis 10; Ver45 = Vermis 4–5*

As a follow-up analysis, we characterized spatial patterns using seed-to-voxel analyses. We tested where seed connectivity was associated with BMI both at an exploratory whole-brain level and within a reward circuit SVC mask (Figure S5; Table S2). Although task-based NAcc connectivity significantly predicted BMI, no corrected whole-brain or reward circuit voxel-wise effects were observed for the NAcc task analysis. The dlPFC seed-to-voxel associations were observed across the whole brain, with no corrected reward circuit SVC effects. The complete voxel-wise results are reported in Table S2.

Finally, to assess whether task- and resting-state FC captured similar or differential spatial patterns, we compared the standardized elastic net weights of ROIs selected in at least one of the modalities for the dlPFC seed. The task-based and resting state weight patterns showed a weak to moderate correspondence (median Pearson *r* = .18 and Spearman *ρ* = .39; Figure S6), with 14 predictors shared between the two models. Of these shared predictors, 92.9% showed weights in the same direction. These findings suggest that task- and resting-state dlPFC FC capture relatively distinct predictive patterns.

## Discussion

BED frequently co-occurs with obesity (35) and has been associated with altered function in brain systems supporting reward processing and cognitive control (16, 17). However, disentangling neural mechanisms related to pathological eating behavior from those associated with obesity remains a major challenge. To this end, FC analyses have been proposed as a promising approach. Yet a robust and replicable fMRI-based neural signature of BED has not been established to date. Using multivariate approaches, we examined FC patterns derived from a priori seed regions during resting state and task fMRI. FC patterns reliably predicted BMI beyond chance, whereas binge eating status could not be predicted using either resting state or task-based FC. Among the examined seed regions, dlPFC connectivity emerged as the most robust predictor of BMI across brain states, while predictive effects of NAcc and ACC connectivity were primarily observed during task performance. Together, these findings suggest that alterations in cognitive control and reward networks are more closely related to obesity than binge eating behavior.

Contrary to our expectations, we found no reliable neural signature of binge eating behavior. Neither multivariate classification analyses nor mass-univariate comparisons of FC maps revealed significant differences between individuals with binge eating behavior and individuals without any history of binge eating. These findings contrast with previous work reporting altered frontostriatal FC in BED. For example, Haynos et al. (21) observed reduced resting state FC between the NAcc and frontal as well as posterior cingulate regions, which was associated with impaired reversal learning and increased binge eating frequency. Here, we were unable to replicate these findings. Instead, an ROI-based analysis suggested a numerically stronger FC among individuals with binge eating behavior in our sample. Differences in sample characteristics, preprocessing choices, or statistical thresholds may partly account for these discrepancies (59). In addition, both studies were characterized by modest sample sizes, which may limit reproducibility and increase sensitivity to sampling variability (60). The absence of statistically significant group differences does not preclude the existence of BED-specific alterations in FC. Instead, our findings suggest that potential BED-related FC differences are smaller than anticipated and potentially context-dependent or overshadowed by obesity-related alterations in brain function.

In contrast to binge eating status, BMI could be reliably predicted from FC patterns within cognitive control and reward networks. Among the examined regions, the dlPFC showed the strongest and most consistent predictive value across both resting state and task conditions. As a central node of the frontoparietal control network, the dlPFC is critically involved in executive functions such as inhibitory control (61), self-regulation (62), and the modulation of food craving (63, 64). The importance of the dlPFC for regulating eating behavior is further supported by preclinical work showing that prefrontal projections to the hypothalamus orchestrate cue-potentiated food intake (65). In humans, stimulation of the dlPFC has been shown to reduce food craving (66). In line with this regulatory role, previous studies have often reported reduced dlPFC connectivity in obesity, particularly within frontoparietal networks (24) and lower global brain connectivity (67). At the other end of the spectrum, anorexia nervosa, which is characterized by excessive cognitive control over food intake, has been associated with increased dlPFC recruitment (68, 69). In addition to reduced FC to some brain regions (e.g., putamen and pallidum), we also observed that BMI prediction was driven by increased FC between the dlPFC and other regions of the motivational circuit, such as the ACC and the (70, 71) whereas decreases have been shown in anorexia nervosa (72) and bulimia nervosa (73). This suggests that obesity-related alterations in dlPFC connectivity may not reflect a uniform reduction, but rather a more specific reorganization of network interactions. Guo et al. (74) recently reported increased variability of FC between the limbic and frontoparietal networks in individuals with higher BMI, suggesting greater fluctuations in the functional coupling between networks involved in reward processing and cognitive control. Increased connectivity may reflect compensatory processes or reduced neural efficiency, indicating that greater neural resources are required to maintain cognitive control in individuals with higher BMI.

Task-based connectivity further enhanced BMI prediction for reward- and control-related regions. While dlPFC FC predicted BMI in both brain states, predictive effects of ACC and NAcc FC emerged primarily during task performance. Participants completed a food-bidding task involving food cues (75) that reliably engage reward and cognitive control systems (76). Task states have been shown to enhance individual FC signatures (77) by increasing the ratio of within-subject to between-subject variability (32, 78). In line with this notion, food-related cognitive and motivational engagement during the task may have amplified obesity-related FC differences within reward and control networks (79). At the same time, the slightly reduced predictive strength of dlPFC FC during task performance may indicate that cognitive engagement partially compensates for intrinsic control deficits that are more apparent during resting state conditions.

Several limitations should be considered when interpreting the present findings. First, the fMRI sample consisted only of women, reflecting the higher prevalence of BED in women (2). In addition, binge eating is influenced by ovarian hormones (80), and future studies may account for menstrual cycle phase or hormone levels. Second, although comparable to many fMRI studies in the field, the sample size limits the robustness and generalizability of predictive modeling approaches. Larger datasets will be necessary to improve statistical power and model stability. Third, compared to the online assessment, the BMI range in our sample was limited by scanner constraints, restricting coverage of severe obesity. Finally, the cross-sectional design precludes causal conclusions about whether FC alterations precede or result from weight gain.

In conclusion, the present study demonstrates that FC within reward and cognitive control networks appears to vary primarily as a function of obesity rather than binge eating behavior in women. While BMI could be reliably predicted from FC patterns, we found no robust neural signature of binge eating behavior. These findings highlight the substantial overlap between obesity and binge eating behavior at the neural level and underscore the challenge of disentangling disorder-specific effects from obesity-related alterations. Future studies should therefore employ larger samples, longitudinal designs, and multiple brain states to better characterize individual variability in reward and cognitive control networks and to clarify their role in the development of obesity and disordered eating.

**Table 1.**
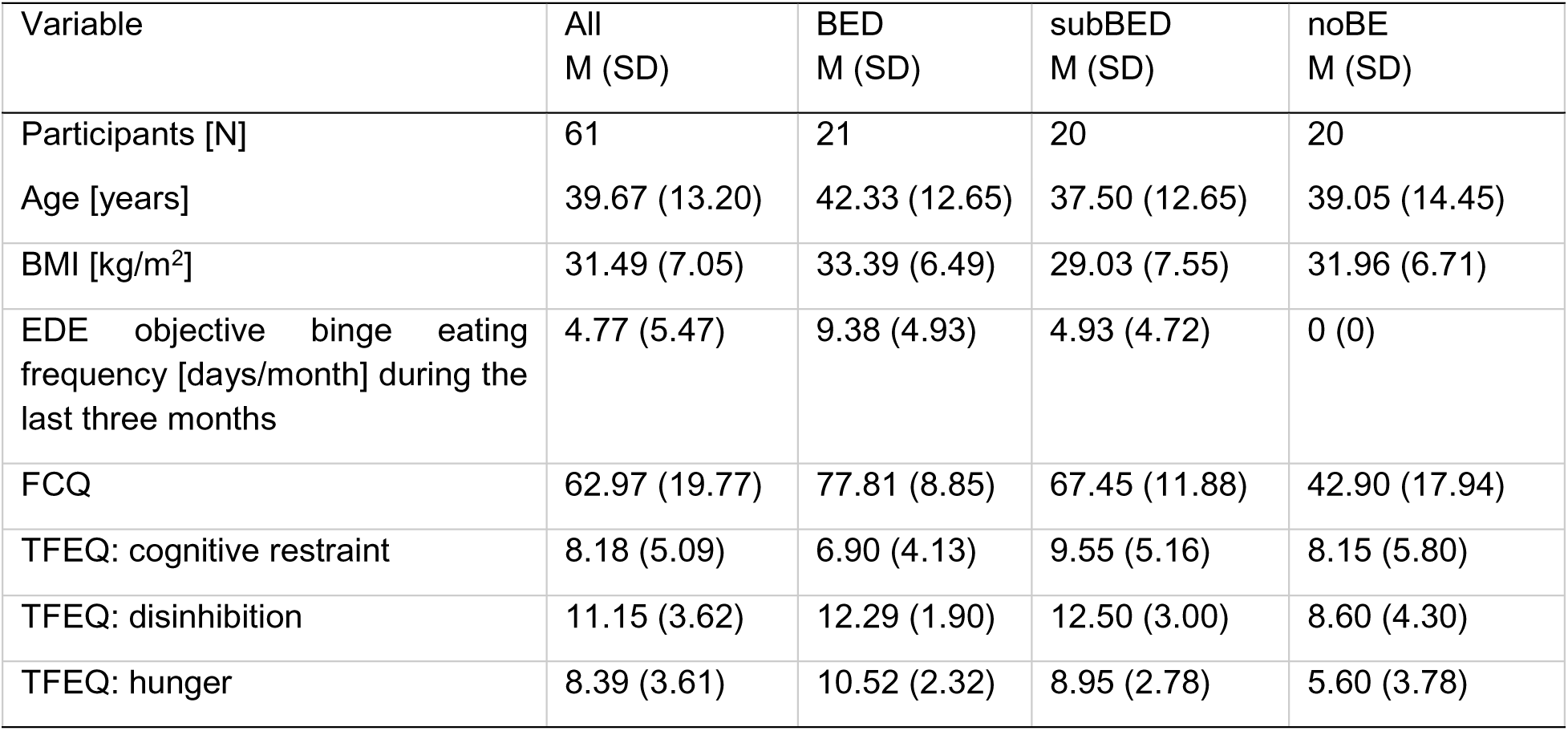
Sample description and summary statistics. M = Mean; SD = Standard Deviation, EDE = Eating Disorder Examination, FCQ = Food Cravings Questionnaire, TFEQ = Three Factor Eating Questionnaire

## Acknowledgments

We thank the TUE002 study team for help with data acquisition. The study was supported by the Else Kröner-Fresenius Stiftung grants 2017_A67 and 2024_EKEA.149 and the Deutsche Forschungsgemeinschaft (DFG, German Research Foundation), grants KR 4555/7-1; KR 4555/9-1; and KR 4555/10-1.

## Author Contributions (CRediT)

**Antonia S. Schlaich:** Formal analysis, Writing – original draft, Writing – review & editing. **María Berjano Torrado:** Formal analysis, Visualization, Software, Writing – original draft, Writing – review & editing. **Anne Kühnel:** Methodology, Software, Data curation, Validation, Visualization, Writing – review & editing. **Jennifer Svaldi:** Conceptualization, Resources, Funding acquisition. **Nils B. Kroemer:** Conceptualization, Resources, Methodology, Supervision, Funding acquisition, Project administration, Visualization, Writing – review & editing.

## Data availability

Group-level statistics are available online at https://identifiers.org/neurovault.collection:17852. Individual connectivity vectors will be shared upon publication. All other data produced in the present study are available upon reasonable request to the authors.

