## Supporting Information for "Functional connectivity of reward and cognitive control networks predicts body mass index but not binge eating disorder in women"

### Supplementary Information

#### fMRI data preprocessing

Preprocessing of resting state fMRI data was performed using fMRIPrep 20.1.1 (1), which is based on Nipype 1.4.1 (2, 3). The T1-weighted (T1w) images were corrected for intensity non-uniformity (INU) with N4BiasFieldCorrection (4) and skull-stripped with a Nipype implementation of the antsBrainExtraction.sh workflow (from ANTs), using OASIS30ANTs as target template. Brain tissue segmentation of cerebrospinal fluid (CSF), white-matter (WM) and gray-matter (GM) was performed on the brain-extracted T1w using fast (FSL 5.0.9, (5)). Brain surfaces were reconstructed using recon-all (FreeSurfer 6.0.1, (6)), and the brain mask estimated previously was refined with a custom variation of the method to reconcile ANTs-derived and FreeSurfer-derived segmentations of the cortical gray-matter of Mindboggle (7). Volume-based spatial normalization to the ICBM 152 Nonlinear Asymmetrical template version 2009c (8) was performed through nonlinear registration with antsRegistration (ANTs 2.2.0), using brain-extracted versions of T1w volume and template.

Correction of distortion was performed using field maps processed with FUGUE (9) with the following program of co-registration to the corresponding T1w using boundary-based registration (10) by BBREGISTER (FreeSurfer). Co-registration was configured with six degrees of freedom. Head-motion parameters with respect to the BOLD reference (transformation matrices and six corresponding rotation and translation parameters) are estimated before any spatiotemporal filtering using MCFLIRT (FSL 5.0.9, (11)). BOLD runs were slice-time corrected using 3dTshift from AFNI (12). BOLD-to-T1w transformation, T1w-to-template (MNI) wrap, field distortion correcting warp, and motion correcting transformations were concatenated and applied in a single step using antsApplyTransforms based on Lanczos interpolation (13). Physiological noise regressors were extracted using Nilearn by calculating the average signal inside anatomically derived CSF and WM masks across time. Framewise displacement of each functional run was calculated using Nipype (14).

Further preprocessing of the functional data was performed using CONN (15) release 22.a and SPM (16) release 12.7771. The functional data were smoothed with a Gaussian kernel of 6 mm full-width half maximum (FWHM). In addition, functional data were denoised using a standard denoising pipeline (17) including white matter timeseries (5 CompCor noise components), CSF timeseries (5 CompCor noise components), six motion parameters and their first order derivatives (18), outlier scans (14), session effects, and linear and quadratic trends within each functional run. This was followed by bandpass frequency filtering of the BOLD timeseries (19) between 0.01 Hz and 0.1 Hz. CompCor (20, 21) noise components within white matter and CSF were estimated by computing the average BOLD signal as well as the largest principal components orthogonal to the BOLD average, motion parameters, and outlier scans within each subject's eroded segmentation masks (22).

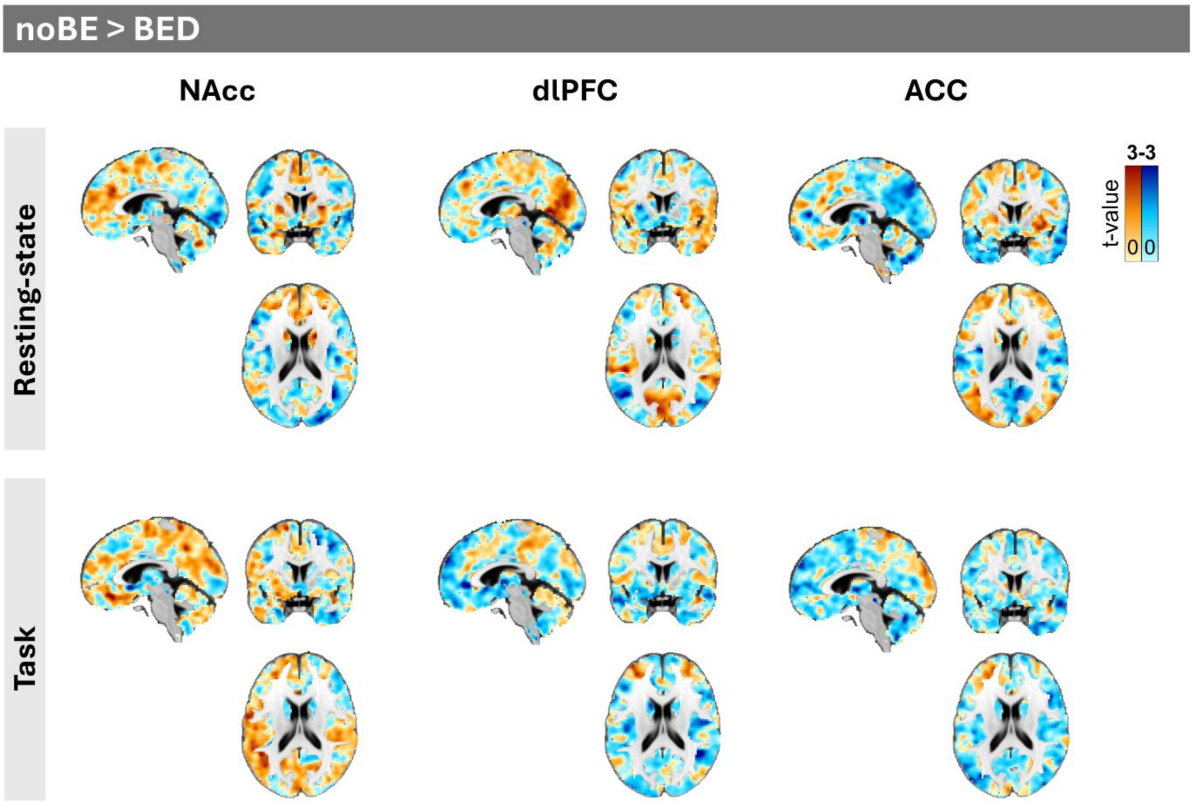


**Figure S1. FC contrast maps between noBE and BED during resting state and task (unthresholded).** The color scale represents t-values, with warm colors indicating stronger functional connectivity in the noBE group compared to the BED group, and cool colors indicating stronger connectivity in the BED group compared to the noBE group. The scale ranges from -3 to 3, with values closer to zero indicating smaller differences between groups.

| Region | Study | MNI Coordinates (x, y, z) | Cluster Size (voxels) | T value | p value |
|---|---|---|---|---|---|
| L SFG (BA 9) | Haynos et al. (2021) | -6, 52, 43 | 963 | 6.09 | <0.001 |
| | Current | -6, 52, 46 | 20 | 0.00 | 0.834 |

| | | | | | |
|---|---|---|---|---|---|
| L PCC (BA 23) | Haynos et al. (2021) | -2, -54, 18 | 269 | 3.88 | <0.001 |
| | Current | 0, -52, 16 | 27 | -0.64 | 0.881 |

**Table S1. Non-replication of FC differences between healthy and BED individuals found by Haynos et al. (2021).** The table shows the results of the NAcc seed FC contrast noBE > BED after applying a small volume correction using the combined mask of the two ROIs reported by Haynos et al. (2021).

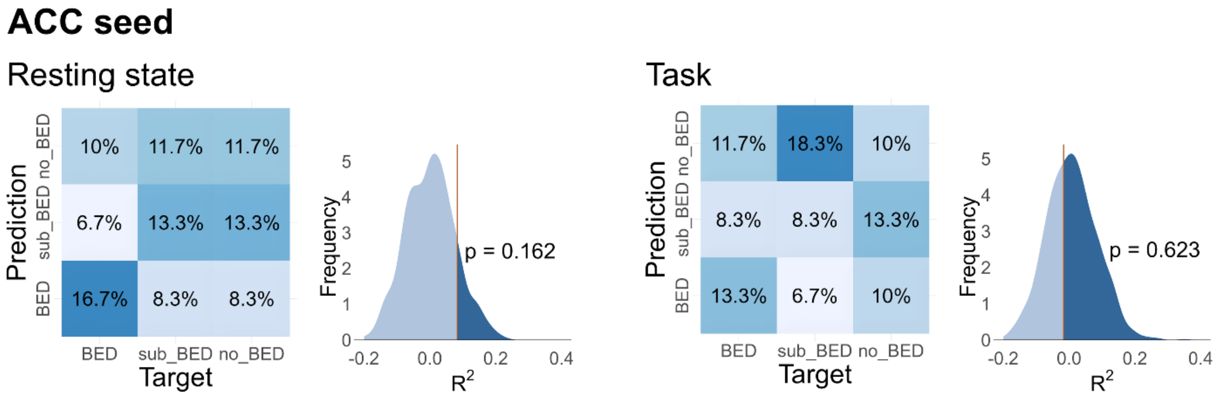


**Figure S2. BED status classification accuracy based on ACC seed-to-ROI FC for resting state and task fMRI.** For each condition, a multiclass confusion matrix shows the normalized proportion of subjects in each observed group (columns) classified into each predicted group (rows). Additionally, the distribution of the classification accuracy generated by permutation testing is shown, with a vertical line representing the observed data test statistic. The median accuracy of the permutation test was used as a comparison baseline (1,000 iterations).

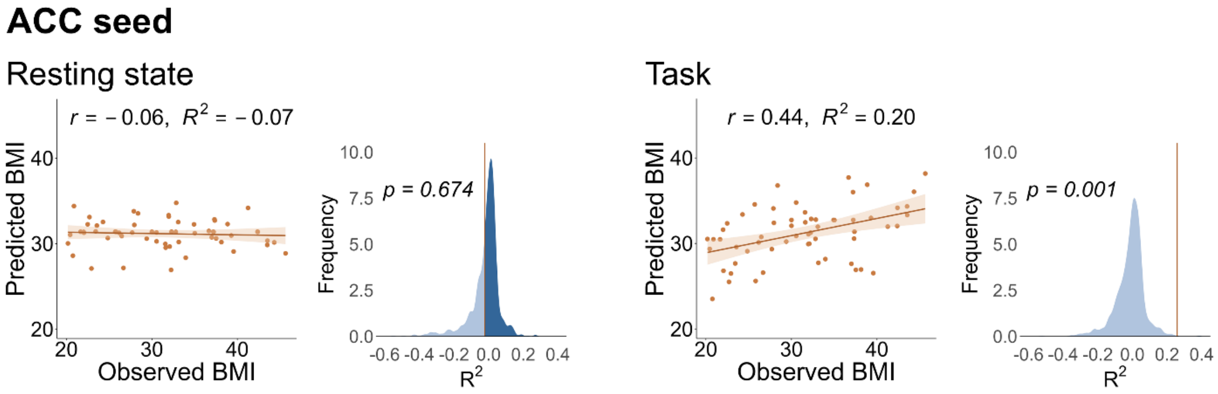


**Figure S3. BMI prediction accuracy based on ACC seed-to-ROI FC for resting state and task fMRI.** For each condition, a scatter plot displays the relationship between observed BMI and predicted BMI, with fitted regression lines and shaded confidence intervals. Additionally, a distribution of the coefficient of determination (R-squared measure of goodness of fit) generated by permutation testing is shown, with a vertical line representing the observed data test statistic. The median coefficient of determination of the permutation test was used as a comparison baseline (i.e., $R^2 = 0$; 1,000 iterations).

ACC-based FC patterns significantly predicted BMI during the task condition ($p = 0.001$, $R^2 = 0.258$), whereas resting-state ACC FC did not ($p = 0.674$, $R^2 = -0.022$; Fig. S3). The task-based elastic net model indicated that stronger ACC connectivity with the caudate, NAcc, and midfrontal regions contributed positively to BMI prediction (Figure S4). Although resting-state ACC-to-ROI connectivity did not significantly predict BMI, resting-state ACC seed-to-voxel analyses revealed peak-level reward circuit SVC effects in the left putamen and caudate ($p_{FWE\text{-}corr}$ = 0.020 and 0.021, respectively). During the task condition, ACC seed-to-voxel effects were observed primarily in posterior occipital regions at the whole-brain cluster level ($p_{FWE\text{-}corr} < 0.001$) rather than in reward circuit regions.

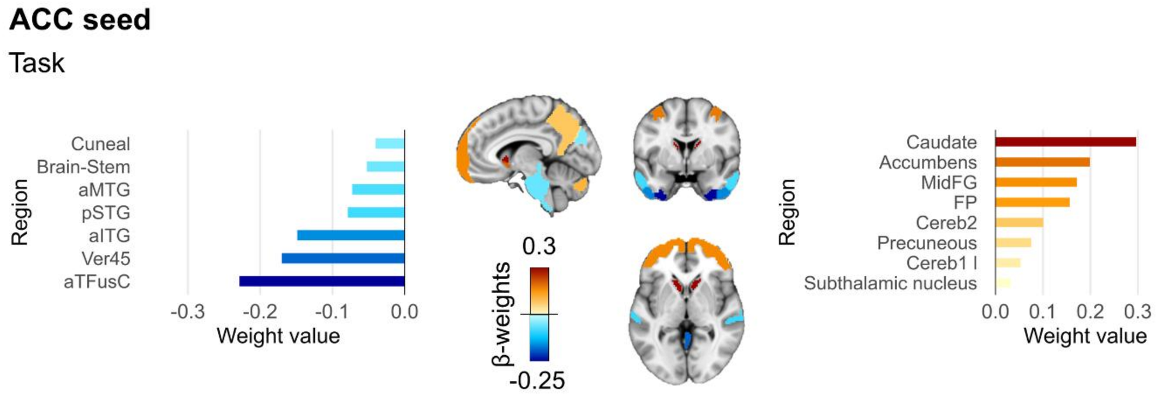


**Figure S4. BMI prediction weight maps of ACC seed during task.** β-weights from the BMI prediction model for ACC seed connectivity during task. Positive weights indicate connections for which higher FC contributed to higher predicted BMI, whereas negative weights indicate connections for which lower FC contributed to higher predicted BMI.

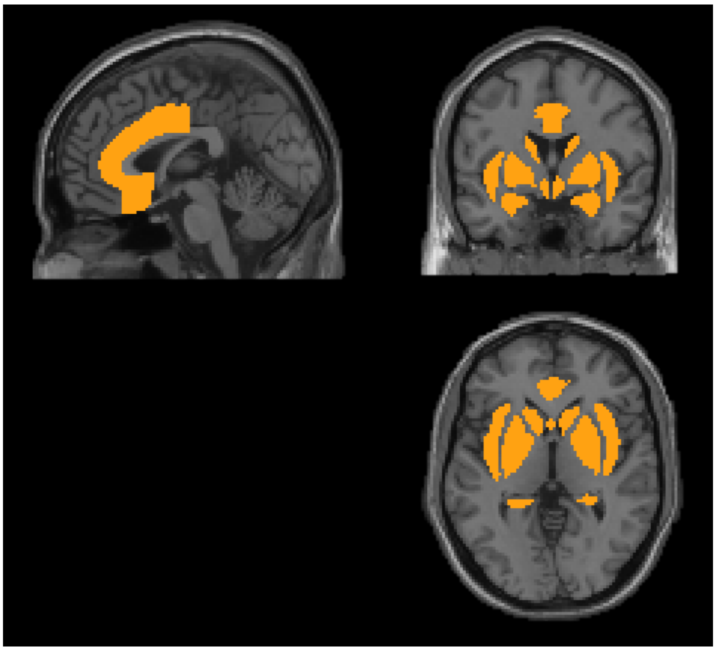

**Figure S5. Binary reward circuit mask used for small-volume correction.** The reward circuit mask was created from the Harvard-Oxford atlas by selecting reward-related regions. Specifically, the voxels with the following atlas labels, bilateral when available, were retained and all other voxels were excluded: insular cortex, subcallosal cortex, anterior division of the cingulate gyrus, caudate, putamen, pallidum, hippocampus, amygdala, substantia nigra pars reticularis, hypothalamus, accumbens, substantia nigra pars compacta, ventral tegmental area, and ventral pallidum.

| Seed | Condition | Search space | Cluster $p_{FWE-corr}$ | k | Region | Hemisphere | Peak $p_{FWE-corr}$ | t | Peak x | Peak y | Peak z |
|---|---|---|---|---|---|---|---|---|---|---|---|
| NAcc | Rest | Whole brain | 0.005 | 164 | midFG | Left | n.s. | 4.10 | -36 | 28 | 40 |
| | | Reward circuit | n.s. | 39 | ACC | Left | 0.042 | 4.75 | -2 | -2 | 38 |
| | Task | Whole brain | n.s. | - | - | - | n.s. | - | - | - | - |
| | | Reward circuit | n.s. | - | - | - | n.s. | - | - | - | - |
| dlPFC | Rest | Whole brain | 0.001 | 226 | PostCG | Left | n.s. | 4.86 | -30 | -36 | 52 |
| | | | 0.01 | 156 | PreCG | Left | n.s. | 4.84 | -48 | -4 | 58 |
| | | | 0.002 | 207 | sLOC | Left | n.s. | 4.28 | -26 | -88 | 40 |
| | | Reward circuit | n.s. | - | - | - | n.s. | - | - | - | - |
| | Task | Whole brain | 0.036 | 150 | PreCG | Right | n.s. | 4.45 | 30 | -14 | 74 |
| | | | 0.019 | 174 | SMA | Right | n.s. | 3.98 | 2 | -6 | 56 |
| | | | | | SMA | Left | n.s. | 3.93 | -6 | -2 | 60 |
| | | | 0.024 | 165 | FP | Left | 0.014 | -5.9 | -34 | 44 | -18 |
| | | Reward circuit | n.s. | - | - | - | n.s. | - | - | - | - |
| ACC | Rest | Whole brain | 0.02 | 138 | CerebWM | Right | n.s. | 4.28 | 14 | 12 | -10 |
| | | | | | Putamen | Right | n.s. | 4.27 | 28 | 14 | -2 |
| | | | | | Putamen | Right | n.s. | 4.12 | 14 | 12 | -10 |
| | | | 0.004 | 187 | iLOC | Left | n.s. | -5.24 | -42 | -64 | 10 |
| | | | 0.029 | 127 | iLOC | Right | n.s. | -4.08 | 42 | -68 | 10 |
| | | Reward circuit | 0.024 | 68 | Putamen | Left | 0.02 | 4.97 | -24 | 8 | -8 |
| | | | n.s. | - | Caudate | Left | 0.021 | 4.96 | -10 | 2 | 10 |
| | | | 0.011 | 88 | Putamen | Right | n.s. | 4.21 | 26 | 14 | -4 |
| | Task | Whole brain | <0.001 | 331 | Cereb1 | Left | 0.04 | 5.58 | -18 | -80 | -20 |
| | | | | | OFusG | Left | n.s. | 4.21 | -38 | -68 | -18 |
| | | | <0.001 | 374 | sLOC | Left | n.s. | 4.95 | -32 | -88 | 10 |
| | | | | | OP | Left | n.s. | 4.19 | -18 | -102 | 16 |
| | | | | | iLOC | Left | n.s. | 3.93 | -42 | -88 | 8 |
| | | | <0.001 | 325 | OP | Right | n.s. | 4.73 | 20 | -94 | 6 |

| | | | | | | | | | | |
|---|---|---|---|---|---|---|---|---|---|---|
| | | <0.001 | 486 | Cereb1 | Right | n.s. | 4.46 | 24 | -82 | -20 |
| | | | | Cereb6 | Right | n.s. | 4.37 | 14 | -80 | -18 |
| | | | | iLOC | Right | n.s. | 4.02 | 44 | -74 | -10 |
| | | n.s. | - | HG | Left | 0.046 | -5.53 | -42 | -22 | 8 |
| | Reward circuit | n.s. | - | - | - | n.s. | - | - | - | - |

**Table S2. Whole-brain and reward circuit small-volume correction seed-to-voxel BMI associations per seed and condition.** Cluster $p_{FWE\text{-}corr}$ and peak $p_{FWE\text{-}corr}$ indicate cluster-level and peak-level FWE-corrected significance for local maxima, respectively. Multiple rows under the same cluster indicate local maxima within that cluster. Local maxima are reported for anatomical localization and are not interpreted as peak-level significant unless peak $p_{FWE\text{-}corr} < 0.05$. Peak t-values are reported as absolute values. Coordinates are in MNI space. *k*: cluster size; *n.s.*: no corrected suprathreshold effect. *ACC = Anterior cingulate cortex; Caudate = Caudate nucleus; Cereb1 = Lobule I of the cerebellum; Cereb6 = Lobule VI of the cerebellum; CerebWM = Cerebellar white matter; dlPFC = Dorsolateral prefrontal cortex; FP = Frontal pole; HG = Heschl's gyrus; iLOC = Inferior lateral occipital cortex; midFG = Middle frontal gyrus; NAcc = Nucleus accumbens; OFusG = Occipital fusiform gyrus; OP = Occipital pole; PostCG = Postcentral gyrus; PreCG = Precentral gyrus; Putamen = Putamen; SMA = Supplementary motor area; sLOC = Superior lateral occipital cortex*

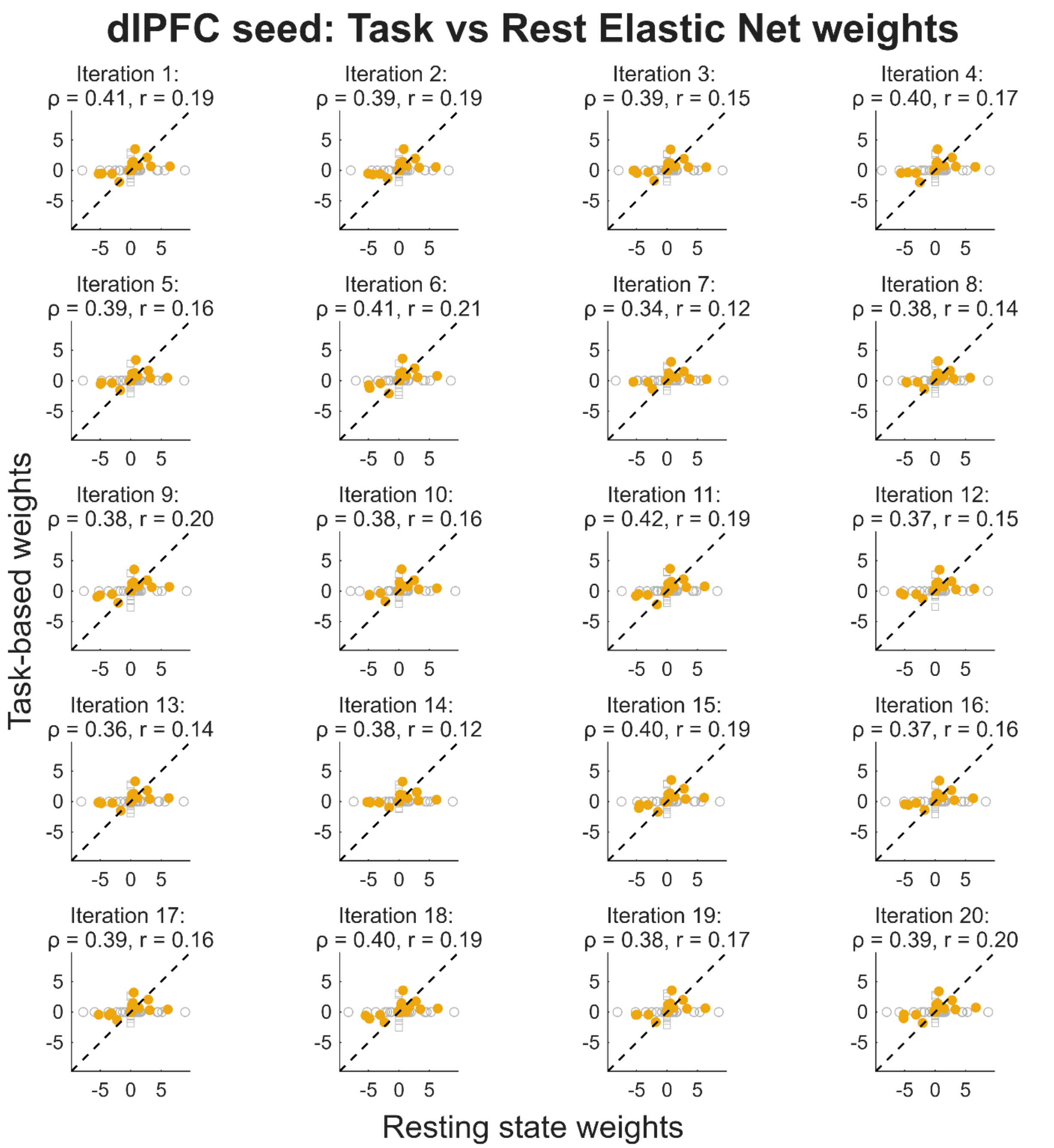


**Figure S6. Weak to moderate similarity of dlPFC FC predictor weights between task-based and resting state models.** Each point represents an FC β-weight, classified according to whether they were selected by both models (filled dots) or by the resting state or task model only. The dashed diagonal indicates identity line. Across 20 iterations, the models showed moderate correspondence in weight ranking (median Spearman $\rho = 0.39$), weak correspondence regarding the absolute weight magnitude (median Pearson $r = 0.167$), and consistent weight direction for the shared predictors (median 92.9%).
